# A novel web-based 24-hour dietary recall tool in line with the Nova food processing classification: description and evaluation

**DOI:** 10.1101/2023.04.28.23289211

**Authors:** Daniela Neri, Kamila Tiemann Gabe, Caroline Dos Santos Costa, Euridice Martinez Steele, Fernanda Rauber, Dirce Maria Marchioni, Maria Laura Louzada, Renata Bertazzi Levy, Carlos Augusto Monteiro

## Abstract

**Objective:** This paper describes the first web-based self-completed 24-hour recall designed to categorize food intake according to Nova groups - Nova24h – and its agreement with a reference tool in estimating the dietary relative contribution of the four Nova food groups (% of total energy intake).

**Design:** Comparisons of estimates of dietary relative contributions of Nova groups obtained by Nova24h and one standard interviewer-led 24-hour recall.

**Setting:** Nationwide adult cohort study in Brazil.

**Participants:** The subjects were 186 participants of the NutriNet Brasil Cohort Study (n=186).

**Results:** No statistically significant differences were observed between the Nova24h and the reference tool mean contributions of unprocessed or minimally processed foods (52.3% vs. 52.6%), processed culinary ingredients (11.6% vs. 11.9%), processed foods (17.1% vs. 14.7%) and ultra-processed foods (19.0% vs. 20.9%). Intraclass correlation coefficients between individual estimates obtained for each Nova group showed moderate to good agreement (0.54-0.78). Substantial or almost perfect agreement between the tools was seen regarding the ability to rank participants according to quintiles of contribution of each Nova group (PABAK 0.69-0.81).

**Conclusions:** Nova24h is a suitable tool for estimating the dietary relative energy contribution of Nova food groups in the NutriNet Brasil cohort. New studies are necessary to verify its adequacy in other populations.

## INTRODUCTION

The Nova food classification is a system that categorizes foods based on the extent and purpose of food processing they undergo^(1)^. Many studies worldwide have used the Nova system to assess diet and health relationships^(2-7)^ nurturing its further use as a framework for national dietary guidelines and dietary guidance from national and international health associations^(8)^. This growing interest in food processing has prompted researchers to explore methods for measuring the extent of processing of dietary data.

Some strategies have been proposed to determine the level of processing of foods in large studies and national surveys collected through widely used standard dietary assessment tools, such as interviewer-led 24-recall and food-frequency questionnaires^(9)^, often lacking details about food processing. Additionally, new tools for collecting dietary data specifically designed to discriminate foods according to the level of food processing have been developed^(10,11)^.

The 24-hour multiple-pass dietary recall applied by trained dietitians is considered a reference method among dietary assessment tools for collecting quantitative data regarding both absolute and relative food group or nutrient dietary intakes^(12)^. It captures detailed dietary information, as interviewers ask individuals to recall and inform in detail all foods and drinks they consumed over the last 24 hours^(13)^. However, this tool is often expensive and labor-intensive for researchers, and time-consuming for study participants^(14)^.

Recently developed, validated and available in a few countries, web-based self-completed 24-hour dietary recall tools offer a low-burden and cost-effective alternative for collecting dietary data. These include the INTAKE24^(15)^ and the Oxford WebQ^(16)^ in the UK, the Automated Self-Administered 24-Hour Dietary Assessment Tool (ASA24)^(17)^, available in Australia, Canada and the US, and the tool used by the NutriNet Santé Cohort Study in France^(18)^. These tools can be used in substitution to the 24-hour multiple-pass dietary recall to obtain dietary intake data and estimate energy and nutrients intakes.

The Nova 24-hour dietary recall (Nova24h) is the first web-based self-completed tool that collects 24-hour food intake data in line with the Nova system. It automatically classifies every food item into one of the four Nova groups: *unprocessed or minimally processed foods, processed culinary ingredients, processed foods* and *ultra-processed foods*. This tool was developed to be used in the NutriNet Brasil Cohort Study launched in January 2020 to investigate prospective associations between dietary patterns and chronic non-communicable diseases^(19)^.

The objective of the present study is to describe the development of the Nova24h tool and to evaluate its agreement with a standard interviewer-led multiple-pass 24-hour dietary recall in estimating the dietary relative energy contribution of each of the four groups as defined by the Nova classification system among participants of the NutriNet Brasil cohort.

## METHODS

### Development of Nova24h

Nova24h web-based self-completed tool was designed by a team of nutrition epidemiologists from the same research center at the University of Sao Paulo that developed the Nova food classification system. It consists of a series of 395 concatenated close-ended questions about all foods and drinks consumed during the previous day. The food list incorporated into the Nova24h was developed based on foods reported by participants of one nationally representative dietary survey conducted in Brazil in 2008-2009^(20)^.

All questions are answered with touches on the mobile screen or clicks on the computer. To enhance the usability of the program, large radio buttons and simple scrolling fields were included. The format of the questionnaire and the structure of the questions were based on feedback from extensive piloting of the tool by researchers from the Center for Epidemiological Research in Health and Nutrition at the University of Sao Paulo. The Nova24h estimated completion time is about 15 minutes.

Before initiating the recall, participants are provided with brief instructions on how to complete the Nova24h questionnaire. They are then asked a question related to food restrictions or special diet (e.g., lactose-, gluten-, and/or red meat-free, vegetarian, or vegan) and redirected to the questions about food items consumed in the previous day as described in Figure 1. Participants are presented with 57 ‘yes/no’ key questions about commonly consumed foods and drinks (e.g., “Did you eat fish yesterday?”). For positive answers, subsequent questions are prompted to specify the type (e.g., fresh fishes such as salmon, tuna, sardine, hake and tilapia, and salted fishes, such as cod or canned fish) and amount consumed of each selected food item (e.g., number of fish filets consumed) (n=190).

**Figure 1.**
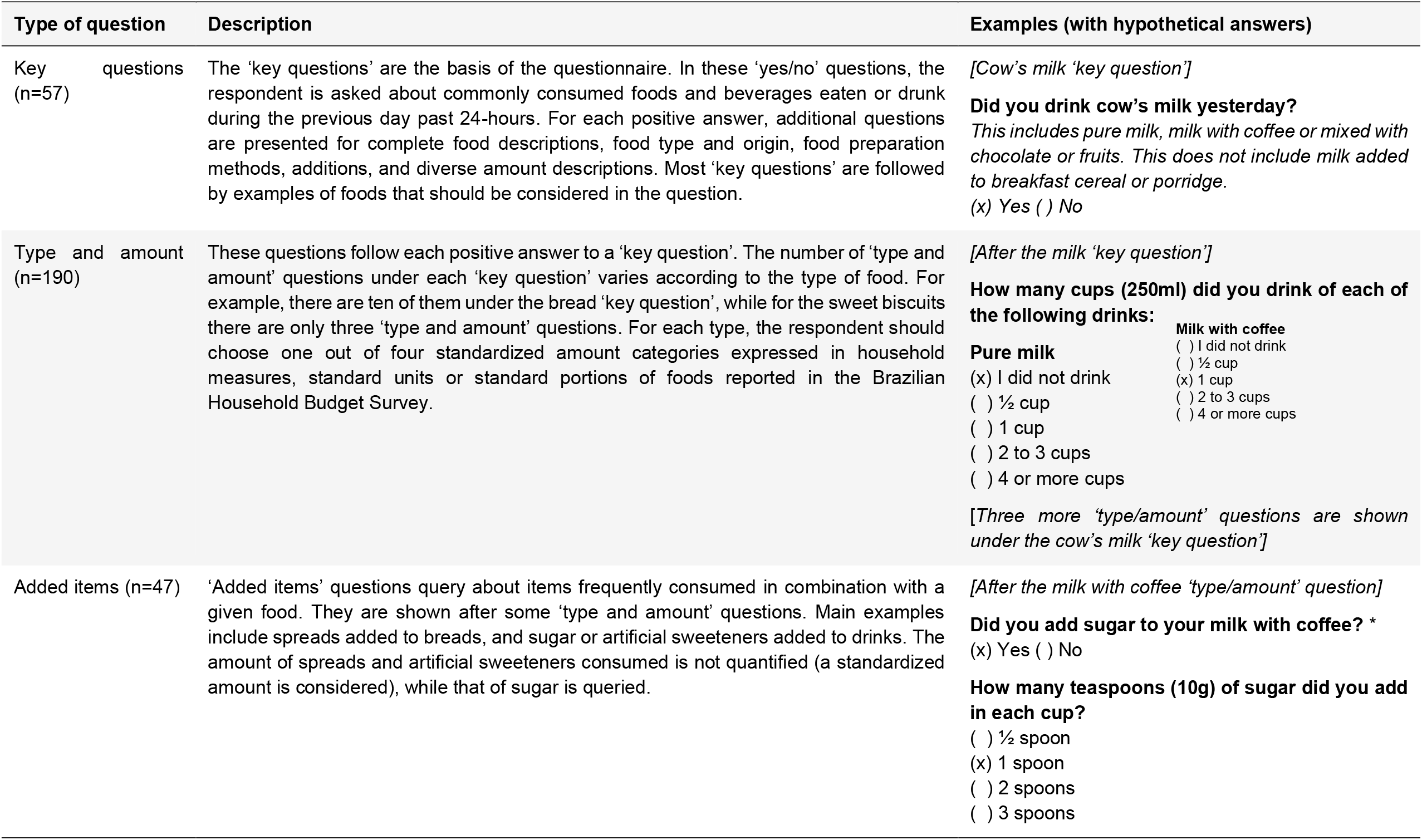
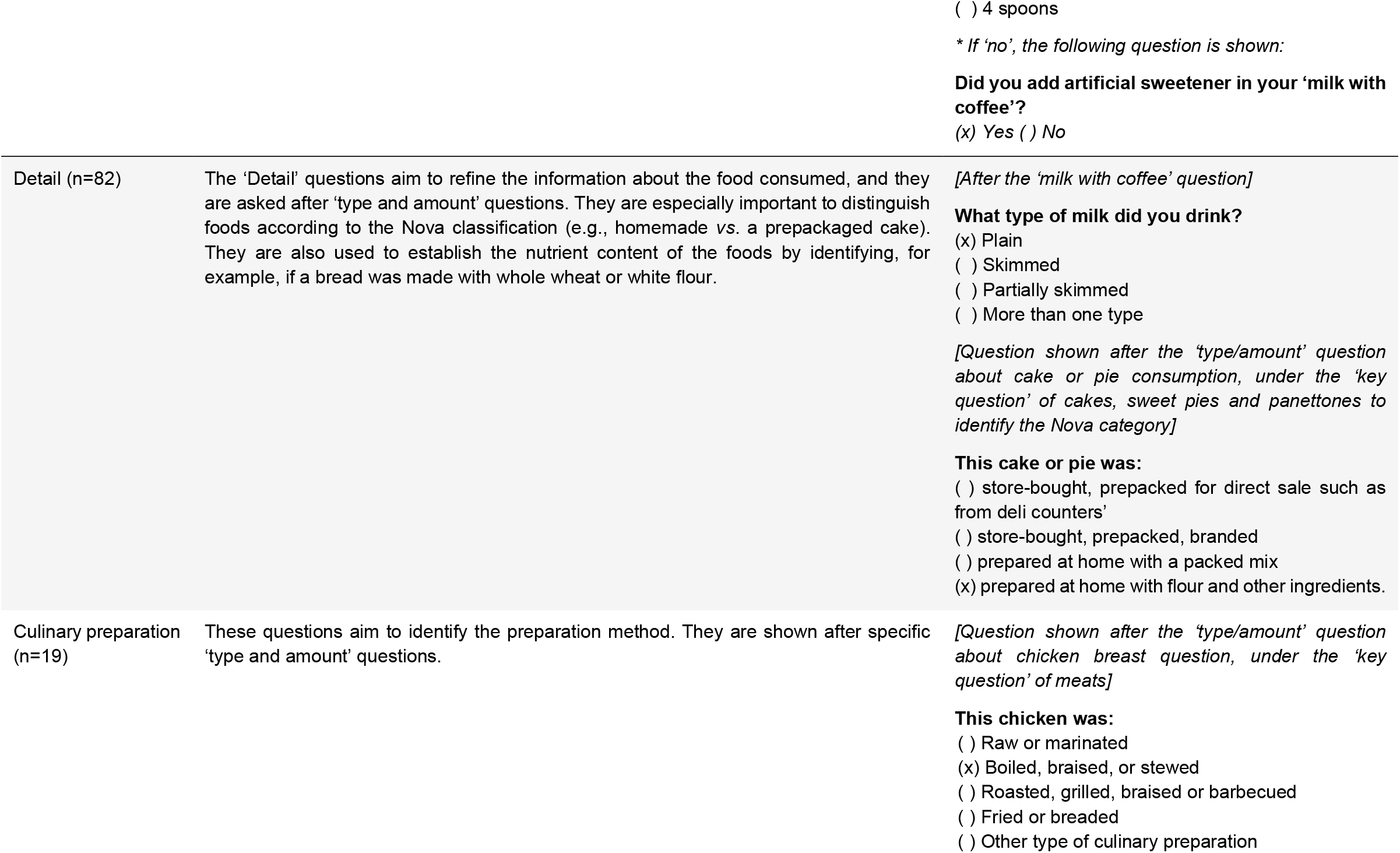
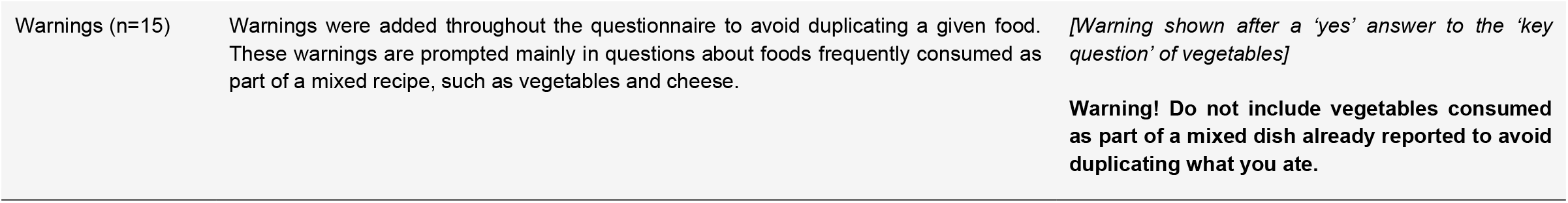
Structure of the Nova24h tool.

The questions were designed to reduce respondent burden by optimizing the number of food items covered by each question. In this sense, each question may refer to one (e.g. cereal bar) or more than one related individual items/culinary preparations (e.g. 1. fruit compote, guava paste, pumpkin jam or marmalade; 2. pudding, manjar or mousse; 3. coxinha, pie, sfiha and kebab). Amounts are reported using standardized categories, including portions of commonly used household measures (e.g., bowls and spoons) and standard units (e.g., an apple, a can)^(20)^.

Certain ‘type and amount’ answers also prompt questions about added items (e.g., sugar added to coffee) (n=47), preparation method in the case of home-made dishes (e.g., raw/marinated, cooked, sautéed or stewed, roasted, grilled, barbecued, fried, breaded, in case of fish) (n=19) and further details to refine the information about the food consumed (e.g., homemade or purchased ready-to-eat, in case of cakes) (n=82).

To avoid the problem of respondents recording the same food item more than once under different questions, warnings were added throughout the recall process (n = 15). For instance, the question “Did you drink milk yesterday?” is followed by the warning: *Attention! DO NOT include milk added to porridge or breakfast cereal here so as not to duplicate what you ate. If that’s the case, change your answer above to “no”*. To aid data interpretation, after completing the recall, participants are asked a short series of questions (n = 3) to determine whether the overall intake was representative of a typical day.

A description of the food items and all their possible variations within each of the 57 key-questions is presented in the Supplementary Table S1. A total of 526 food items capture all possible combinations of responses to questions within each of the 57 key-questions, including 347 individual or grouped items (e.g. ‘whole milk’ or ‘squash, zucchini or eggplant’) and 179 culinary preparations, which are subsequently disaggregated into underlying ingredients (e.g. “cooked rice” is disaggregated into: rice, oil, onion, garlic and salt). Standardized recipes from the *Tabela Brasileira de Composição de Alimentos* 7.0^(21)^ (TBCA - Brazilian Table of Food Composition) were the primary source used for the disaggregation. When a standardized recipe for some specific dish was not available in the TBCA (n=10), an adapted TBCA recipe was used to attribute ingredients and their respective proportions.

The nutritional composition of each individual or grouped food items and ingredients from the culinary preparations was estimated using food codes from the TBCA. Food codes from the United States Department of Agriculture^(22)^ National Nutrient Database were used to code foods with no match with the TBCA (11 out of 526). For grouped food items, the food code representing the most consumed food (according to the national survey^(20)^) was used (e.g. “zucchini” food code was used for coding “squash, zucchini or egg-plant”).

A three-stage process was undertaken to classify the 347 food items and ingredients of 179 culinary preparations according to the extent and purpose of industrial food processing as established by the Nova food classification system. First, two researchers working independently (E.M.S and C.S.C) assigned food items and ingredients to one of four mutually exclusive Nova groups and subgroups (Supplementary Table S2). Second, Nova food groups and subgroups data were reviewed independently by two separate researchers (D.N and K.G). Food items and ingredients for which there was consensus in the categorization among all researchers were assigned to their Nova group and subgroup. Food items with disagreement in categorization between any two researchers were shortlisted and flagged for further scrutiny. At stage three, an expert panel of two nutrition epidemiologists (R.B.L and M.L.L) with substantial experience working with the dietary intake in the national dietary survey was convened to review, discuss and reach an agreement about the categorization of the short-listed products.

A data set in a long format including the 526 food items and the underlying ingredients of culinary recipes, as well as their NOVA classification, food codes and nutritional composition, was built into the system. Using this matrix, Nova24h automatically generates an output informing all the foods and amounts consumed by the respondents with their respective nutritional content and classification according to Nova. Though the Nova24h was designed to estimate the energy and nutrient contents of the diet, its ability to do so in comparison to a standard tool has not yet been tested.

### Evaluation of the agreement between Nova24h and the standard method

#### Study sample

All participants of the NutriNet Brasil Cohort Study who completed the Nova24h tool between September 18th and October 16th of 2020 were consulted in the online platform about whether they would accept to participate in the agreement study. Among those who agreed to participate (nearly 3/4 of participants), a sample of 186 participants, selected to mimic the demographic distribution (age, sex, and region of residence) of the total adult Brazilian population, was studied. A sample size of at least 152 is required for reaching 80% power in detecting even weak agreements (Intraclass correlation coefficient = 0.2) between two observations per subject^(23)^.

#### Data collection

Following completion of the Nova24h recall, the selected sample was contacted over the phone on the same day to respond to the reference method – the interviewer-led multiple-pass 24-hour dietary recall^(13)^. Eighty-five percent of participants completed the two recalls on weekdays and 15% on weekend days.

The multiple-pass interviews were carried out using the Brazilian version of the GloboDiet software^(24)^ by two dietitians skilled in the use of this tool. The dietitians were trained to pay particular attention to food intake information needed to capture the level of processing of food items. This included specific information on the preparation or processing of certain food items (e.g., home-made from scratch or ready-to-eat products), brand names of packaged products (for branded breakfast cereal and breads, for instance), the place of preparation (at restaurant, street-food, take-away places), as well as the method of preparation of mixed dishes and the types of ingredients used (e.g., from scratch with fresh ingredients or pre-made and frozen). The dietitians were blinded to what the participants had entered in the Nova24h. All food items from the GloboDiet database were coded into food codes and subsequently classified according to Nova by the same researchers (E.M.S and C.S.C) using the same procedures used in Nova24h. The same TBCA food composition table used in Nova 24h was used to calculate nutrient intakes in the GloboDiet.

#### Data analysis

Standardized procedures were taken to impute or logically calculate estimations. When individuals selected a food item and its amount but did not complete data-fields relating to food type (e.g., plain or skimmed), source (e.g., homemade, or packed) or preparation (e.g., roasted, or fried) (n=23 participants), the amount informed was distributed among all options available for each food, following the distributions observed in the first 27,927 participants of the NutriNet Brasil Cohort Study who had completed one Nova24h. In the current analysis for example, if a participant did not inform the origin of honey bread, 20.4% of the reported amount was considered ‘homemade’, 28% ‘bought at a bakery’ and, 51.6% ‘branded packed’, as these were the proportions reported by the 27,927 participants. Quality control procedures were implemented to verify that appropriate data selection, calculation methods, and data entry were used.

Descriptive statistics including mean values (and standard deviations) and frequency distribution were used to describe sample characteristics. The mean dietary contribution of each Nova group expressed in percentage of total energy intake, with 95% confidence intervals, obtained with Nova24h was compared with the same estimates obtained with the reference-method.

Intraclass correlation coefficients (ICC), derived from two-way mixed-effects models, were used to assess the strength of agreement between the methods in the overall sample. ICC across socio-demographic and weight status categories were assessed as secondary analysis. Values less than 0.50 were interpreted as indicative of *poor* agreement, between 0.50 and 0.75 as *moderate* agreement, between 0.75 and 0.90 as *good* agreement, and values greater than 0.90 as indicative of *excellent* agreement^(25)^.

Finally, to assess whether the methods agree in ranking individuals into the same or adjacent quintiles of consumption of each Nova food group, we divided participants into quintiles of the dietary contribution of each Nova group (% of total energy intake) as estimated by each method.

The percentage classified into the same quintile by both methods was calculated and prevalence-adjusted and bias-adjusted kappa (PABAK) was used to evaluate the level of agreement. Values of PABAK were interpreted as follows: ≤0 *no agreement*, less than 0.20 *none to slight*, 0.21–0.40 *fair*, 0.41–0.60 *moderate*, 0.61–0.80 *substantial*, and 0.81–1.00 indicating *almost perfect*^(26)^.

Comparison between the two methods regarding the dietary energy contribution of subgroups within each Nova group was also performed as secondary analyses. All analyses were conducted using the STATA statistical software package version 15.0.

## RESULTS

Socio-demographic characteristics of the study participants are shown in Table 1. Mean age was 41.3 years, 55% were women, 61% were from the most populous Brazilian regions (Southeast and Northeast) and 95.1% had completed secondary school (46.2%) or college/university (48.9%). The mean BMI of study participants was 26.8 kg/m2; 30% of participants were overweight and 24% obese.

**Table 1.** Socio-demographic and anthropometric characteristics of study participants (n=186)

| Characteristics | Mean ± SD | n (%) |
| --- | --- | --- |
| Age (years) | 41.3 ± 12.0 | |
| Sex |  |  |
| Female |  | 103(55.0) |
| Region |  |  |
| North |  | 16 (8.6) |
| Northeast |  | 53 (28.5) |
| Center-West |  | 22 (11.8) |
| Southeast |  | 60 (32.3) |
| South |  | 35 (18.8) |
| Educational level |  |  |
| Less than elementary |  | 4 (2.2) |
| Elementary |  | 5 (2.7) |
| Secondary |  | 86 (46.2) |
| College/university |  | 91 (48.9) |
| BMI status |  |  |
| < 18.5 kg/m<sup>2</sup> |  | 3 (1.6) |
| 18.5-25 kg/m<sup>2</sup> |  | 82 (44.1) |
| 25-29.9 kg/m<sup>2</sup> |  | 56 (30.1) |
| ≥30 kg/m<sup>2</sup> | | 45 (24.2) |

The contribution of Nova food groups to the total energy intake estimated with Nova24h or with the reference tool (24-hour multiple-pass dietary recall) is shown in Table 2. No statistically significant differences were observed between the Nova24h and the reference tool mean contributions of unprocessed or minimally processed foods (52.3% vs. 52.6%), processed culinary ingredients (11.6% vs. 11.9%), processed foods (17.1% vs. 14.7%) and ultra-processed foods (19.0% vs. 20.9%).

**Table 2.** Dietary contribution (% of total energy intake) of Nova food groups using the Nova24h tool or the interviewer-led 24-hour dietary recall (reference method) (n=186)

| Nova food groups | Nova24h |  |  | Reference method |  |  | Intraclass correlation coefficients |  |  |
| --- | --- | --- | --- | --- | --- | --- | --- | --- | --- |
|  | Mean | 95% CI |  | Mean | 95% CI |  | ICC | 95% CI |  |
| Unprocessed or minimally processed foods | 52.3 | 49.9 | 54.7 | 52.6 | 49.9 | 55.2 | 0.78 | 0.71 | 0.84 |
| Processed culinary ingredients | 11.6 | 10.4 | 12.8 | 11.9 | 10.6 | 13.2 | 0.54 | 0.38 | 0.65 |
| Processed foods | 17.1 | 15.2 | 19.0 | 14.7 | 12.7 | 16.7 | 0.72 | 0.62 | 0.79 |
| Ultra-processed foods | 19.0 | 17.0 | 21.0 | 20.9 | 18.5 | 23.2 | 0.75 | 0.66 | 0.81 |

ICC, Intraclass correlation coefficients

ICC between individual estimates obtained with each tool showed *moderate* agreement for both processed culinary ingredients (0.54; 95 % CI 0.38 – 0.65) and processed foods (0.72; 95 % CI 0.62 - 0.79) and *good* agreement for both unprocessed or minimally processed foods (0.78; 95 % CI 0.71 – 0.84) and ultra-processed foods (0.75; 95 % CI 0.66 - 0.81). ICC did not substantially change across socio-demographic or weight status categories (Supplementary Tables S3 to S6).

The mean dietary contribution of food subgroups within each Nova group estimated with Nova24h or with the reference tool and the corresponding ICC are presented in Supplementary Table S2. *Moderate* or *good* agreement was seen for most subgroups of unprocessed or minimally processed foods, processed culinary ingredients, processed foods and ultra-processed foods.

Table 3 assesses whether the two tools agree in ranking individuals into the same or adjacent quintiles of consumption of each Nova food group and inform the corresponding PABAK. *Substantial* agreement was seen for unprocessed or minimally processed foods, processed culinary ingredients and ultra-processed foods (PABAK of 0.78, 0.69, and 0.77, respectively) and *almost perfect* agreement for processed foods (PABAK of 0.81).

**Table 3.**
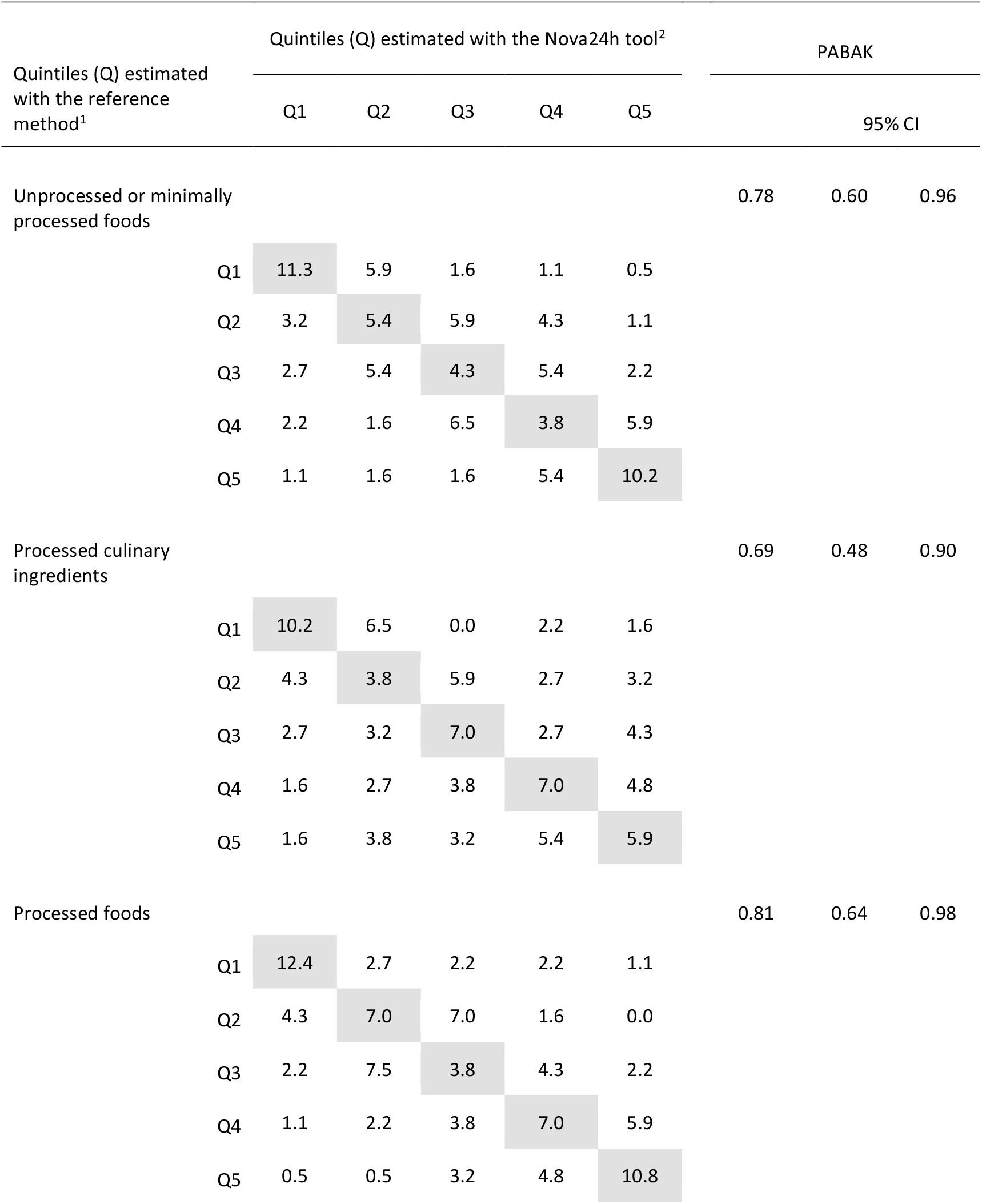
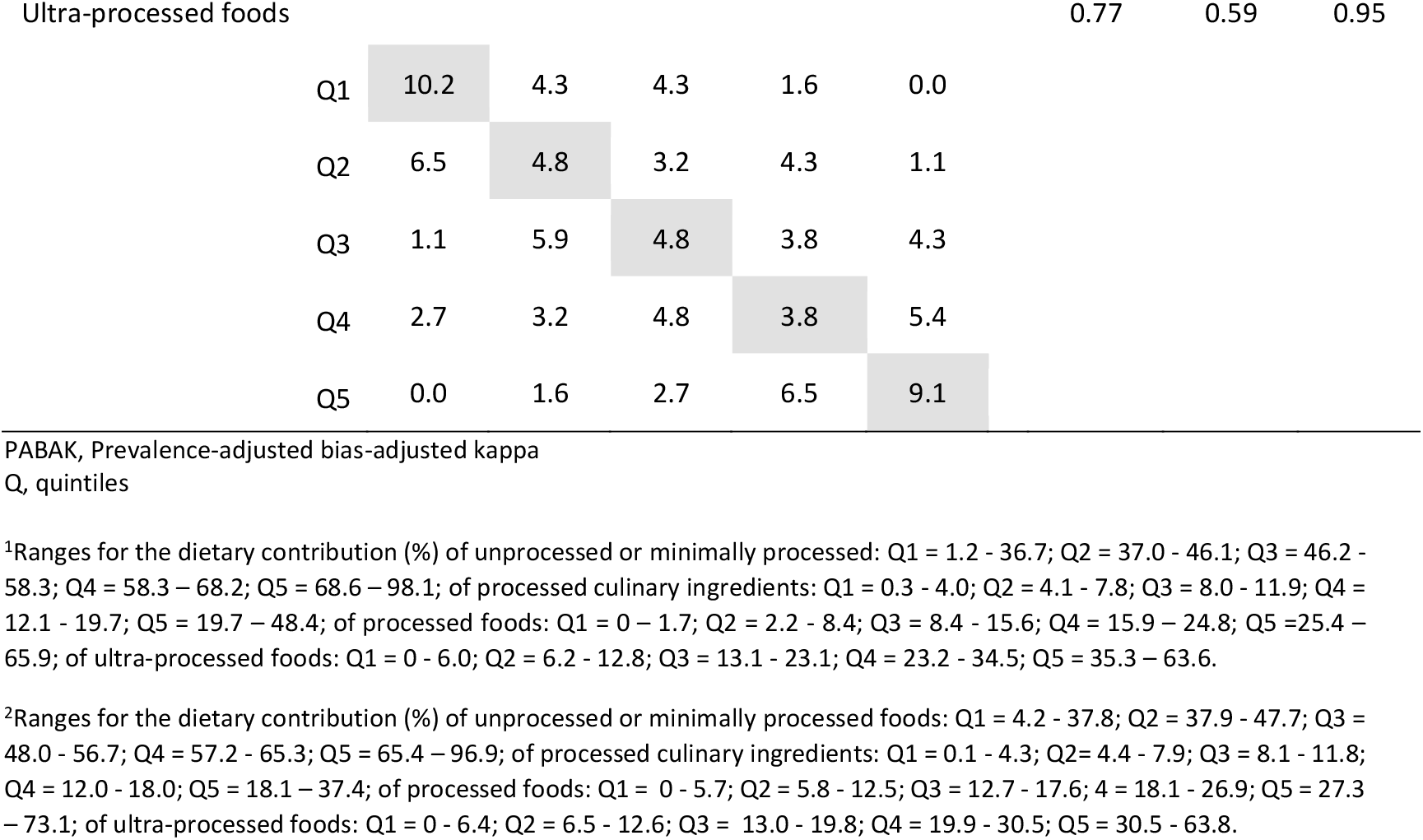
Agreement between participants classification according to quintiles of the dietary energy contribution of each Nova group estimated using the Nova24h and the interviewer-led 24-hour dietary recall (reference tool) (n = 186)

## DISCUSSION

The present study described the first web-based self-completed 24-hour dietary recall tool designed to assess dietary intake in line with the Nova food classification system and evaluated the agreement between this new tool and a reference tool in estimating the dietary contribution of each of the four Nova groups in a sample of participants of the NutriNet Brasil Cohort Study.

We found that the mean dietary energy contribution of each Nova group was almost identical or very close when estimated by the Nova24h or the interviewer-led multiple-pass 24-hour dietary recall tool. The agreement between the two tools in estimating participants’ individual dietary contribution of Nova groups was *moderate* for processed culinary ingredients and processed foods and *good* for unprocessed or minimally processed foods and ultra-processed foods. The agreement to rank participants according to quintiles of each Nova food group consumption was *substantial* for unprocessed or minimally processed foods, processed culinary ingredients and ultra-processed foods and *almost perfect* for processed foods.

The lower agreement between the two tools for processed culinary ingredients (ICC of 0.54 against > 0.70 for the other three Nova groups) is probably explained by the fact that, contrary to the reference tool, Nova24h does not ask participants about oils added to each preparation after they were cooked. This is confirmed by the low ICC regarding vegetable oils shown in Supplementary table S2 (0.31). The initial decision to not include oils added at the table was made to reduce participants’ burden but a new updated version of Nova24h will include this question.

As other web-based self-completed dietary recall tools^(14-18)^, Nova24h has many advantages over interviewer-led recalls. It allows considerable logistic simplification and cost savings; it provides greater flexibility, allowing the subject to complete the recall at any time via a user-friendly interface. Importantly, such advantages may increase participation and retention rates in epidemiological studies. Its main limitation, also shared with similar tools, is the requirement of participants’ literacy and minimum computer skills.

Different from existing web-based self-completed dietary recall tools^(14-18)^, Nova24h was developed to address the need for accurately assessing dietary intake according to food processing levels as defined by the Nova food classification system.

Some study limitations should be noted. Although a sample size of 186 is considered enough for detecting even weak agreements^(23)^, it was not calculated to examine differences according to sociodemographic characteristics or weight status. Though weekend days were underrepresented in this comparative study sample, this will unlikely change the agreement between the tools, since any deviations from true values would probably affect both tools. Also, due to the high schooling levels of the NutriNet Brasil Cohort^(19)^, 95% of the study participants had at least completed high-school education, a condition found only in nearly half of the Brazilian adult population^(27)^. Thus, caution is required when extrapolating the present results to the general Brazilian population or to other Brazilian populations with lower education. Adaptations to the original Nova24h recall are probably necessary for its application in populations with differing dietary patterns from those found in Brazil.

In terms of study strengths, the study design ensured that both tools collected dietary intake data for the same 24 hours and analyzed them using the same food composition table. In addition, the same researchers applied the Nova classification to both sets of data.

Our findings suggest that the low-cost Nova24h may be a suitable method for assessing dietary data according to food processing. Further work will include the evaluation of the performance of this new tool for estimating energy and nutrient intakes.

## CONCLUSION

This study indicates that Nova24h is a suitable tool for assessing dietary relative energy contribution in line with the Nova food system classification among participants of the NutriNet Brasil Cohort Study. New studies are necessary to verify the potential application of Nova24h in other populations.

## Data Availability

All data produced in the present study are available upon reasonable request to the authors

## Abbreviations

G1: unprocessed/minimally processed foods
G2: processed culinary ingredients
G3: processed foods
G4: ultra-processed foods.

## Acknowledgements

The authors wish to acknowledge each of the participants who took part in the study. The authors also gratefully acknowledge the assistance of researchers from the Center for Epidemiological Research in Nutrition and Health for piloting of the Nova24h dietary recall tool.

## Authorship

D. N. designed the tool, trained data collectors, managed the data collection, analyzed, and interpreted the data, and prepared the first draft of the manuscript. K.T.G., C.S.C., E.M.S. and F.R. designed the tool, contributed to the analysis and interpretation of the data, and assisted in writing the paper. D.M.M. trained data collectors, managed the data collection, and reviewed the manuscript. R.B.L. and M.L.L designed the tool and critically reviewed the manuscript. C.A.M. conceived, designed, and supervised the study, interpreted the data, and had primary responsibility for the final content. All the authors approved the manuscript.

## Ethical Standards Disclosure

“This study was conducted according to the guidelines laid down in the Declaration of Helsinki and all procedures involving research study participants were approved by the Human Subjects Research Office at the University of Sao Paulo (USP no. 2.728.201). Written informed consent was obtained from all subjects/patients.

## REFERENCES

1. Monteiro CA, Cannon G, Moubarac JC, et al. (2018) The UN Decade of Nutrition, the NOVA food classification and the trouble with ultra-processing. Public Health Nutr 21, 5–17.

2. Hall KD, Ayuketah A, Brychta R, et al. (2020) Ultra-Processed Diets Cause Excess Calorie Intake and Weight Gain: An Inpatient Randomized Controlled Trial of Ad Libitum Food Intake. Cell Metab 32, 690.

3. Delpino FM, Figueiredo LM, Bielemann RM, et al. (2022) Ultra-processed food and risk of type 2 diabetes: a systematic review and meta-analysis of longitudinal studies. Int J Epidemiol 51, 1120–1141.

4. Pagliai G, Dinu M, Madarena MP, et al. (2021) Consumption of ultra-processed foods and health status: a systematic review and meta-analysis. Br J Nutr 125, 308–318.

5. Suksatan W, Moradi S, Naeini F, et al. (2021) Ultra-Processed Food Consumption and Adult Mortality Risk: A Systematic Review and Dose-Response Meta-Analysis of 207,291 Participants. Nutrients 14, 174.

6. Wang M, Du X, Huang W, et al. (2022) Ultra-processed Foods Consumption Increases the Risk of Hypertension in Adults: A Systematic Review and Meta-Analysis. Am J Hypertens 35, 892–901.

7. Srour B, Kordahi MC, Bonazzi E, et al. Ultra-processed foods and human health: from epidemiological evidence to mechanistic insights. Lancet Gastroenterol Hepatol 7, 1128–1140.

8. Monteiro CA, Astrup A. (2022) Does the concept of “ultra-processed foods” help inform dietary guidelines, beyond conventional classification systems? YES. Am J Clin Nutr 116, 1489–1491.

9. Khandpur N, Rossato S, Drouin-Chartier JP, et al. (2021) Categorising ultra-processed foods in large-scale cohort studies: evidence from the Nurses’ Health Studies, the Health Professionals Follow-up Study, and the Growing Up Today Study. J Nutr Sci 10, e77.

10. Dinu M, Bonaccio M, Martini D, et al. (2021) Reproducibility and validity of a food-frequency questionnaire (NFFQ) to assess food consumption based on the NOVA classification in adults. Int J Food Sci Nutr 72, 861–869.

11. Sarbagili-Shabat C, Zelber-Sagi S, Fliss Isakov N, et al. (2020) Development and validation of processed foods questionnaire (PFQ) in adult inflammatory bowel diseases patients. Eur J Clin Nutr 74, 1653–1660.

12. Dietary Assessment Primer. Principles Underlying Recommendations [Internet]. National Institutes of Health, National Cancer Institute. Available at https://dietassessmentprimer.cancer.gov/ (accessed February 2022).

13. Moshfegh AJ, Rhodes DG, Baer DJ, et al. (2008) The US Department of Agriculture Automated Multiple-Pass Method reduces bias in the collection of energy intakes. Am J Clin Nutr 88,324–32.

14. Park Y, Dodd KW, Kipnis V, et al. (2018) Comparison of self-reported dietary intakes from the Automated Self-Administered 24-h recall, 4-d food records, and food-frequency questionnaires against recovery biomarkers. Am J Clin Nutr 107, 80–93.

15. Bradley J, Simpson E, Poliakov I, et al. (2016) Comparison of INTAKE24 (an online 24-h dietary recall tool) with interviewer-led 24-h recall in 11–24 year-old. Nutrients 8, 358.

16. Liu B, Young H, Crowe FL, et al. (2011) Development and evaluation of the Oxford WebQ, a low-cost, web-based method for assessment of previous 24 h dietary intakes in large-scale prospective studies. Public Health Nutr 14, 1998–2005.

17. Thompson FE, Dixit-Joshi S, Potischman N, et al. (2015) Comparison of Interviewer-Administered and Automated Self-Administered 24-Hour Dietary Recalls in 3 Diverse Integrated Health Systems. Am J Epidemiol 181, 970–8.

18. Touvier M, Kesse-Guyot E, Mejean C, et al. (2011) Comparison between an interactive web-based self-administered 24 h dietary record and an interview by a dietitian for large-scale epidemiological studies. Br J Nutr 105, 1055–64.

19. Steele EM, Rauber F, Costa CDS, et al. (2020) Dietary changes in the NutriNet Brasil cohort during the covid-19 pandemic. Rev Saude Publica 54, 91.

20. Instituto Brasileiro de Geografia e Estatística (2011) Pesquisa de Orçamentos Familiares 2008-2009. Análise do Consumo Alimentar Pessoal no Brasil. Available at https://biblioteca.ibge.gov.br/visualizacao/livros/liv50063.pdf (accessed February 2019).

21. Tabela Brasileira de Composição de Alimentos (TBCA). Universidade de São Paulo (USP). Food Research Center (FoRC). Versão 7.1. São Paulo, 2020. Available at http://www.fcf.usp.br/tbca (accessed October 2020).

22. US Department of Agriculture. Agriculture Research Services. The USDA food and nutrient databases for dietary studies, 4.1-documentation and users guide. Available at http://www.ars.usda.gov/SP2UserFiles/Place/12355000/pdf/fndds_doc.pdf (accessed October 2020).

23. Bujang MA, Baharum N. A simplified guide to determination of sample size requirements for estimating the value of intraclass correlation coefficient: a review. Arch Orofac Sci. 2017;12(1): 1–11.

24. Steluti J, Crispim SP, Araujo MC, et al. (2020) Technology in Health: Brazilian version of the GloboDiet program for dietary intake assessment in epidemiological studies. Rev Bras Epidemiol 23, e200013.

25. Koo TK, Li MY. (2016) A Guideline of Selecting and Reporting Intraclass Correlation Coefficients for Reliability Research. J Chiropr Med 15, 155–163.

26. Landis JR, Koch GG. (1977) The measurement of observer agreement for categorical data. Biometrics 33, 159–74.

27. Instituto Brasileiro de Geografia e Estatística. Coordenação de Trabalho e Rendimento. Título Secundário: Pesquisa Nacional por Amostra de Domicílios Contínua: educação: 2019. Available at https://biblioteca.ibge.gov.br/visualizacao/livros/liv101736_informativo.pdf (accessed July 2022).

