## Supplemental Tables for "A novel web-based 24-hour dietary recall tool in line with the Nova food processing classification: description and evaluation"

**Supplementary Table S1. Description of food items and all their possible variations within each of the 57 key-questions.**

|  | INGLES |
| --- | --- |
| 1 | Plain water; |
| 2 | Coffee with and without added sugar or artificial sweetener; |
| 3 | Milk or beverages alternatives, with and without added sugar, in their classical, diet or light forms; |
| 4 | Milk, including pure milk, milk with coffee, chocolate, and fruit, with and without added sugar or artificial sweetener, in whole, skim or low-fat versions; |
| 5 | Soft drinks, in their classical, diet, light or zero sugar forms; |
| 6 | Freshly squeezed fruit juice made from fruit or pulp, with and without added sugar or artificial sweetener; |
| 7 | Packaged fruit juice, including “100% fruit juice” or brands like “Tang”, “Maguary” or others, with and without added sugar or artificial sweetener; |
| 8 | Plain and flavored yogurt, which can be whole or low-fat, with and without added sugar or artificial sweetener; |
| 9 | Homemade tea, with and without added sugar or artificial sweetener; |
| 10 | Other packaged beverages, including iced tea, sports or energy drinks, or nutritional supplements. |
| 11 | Alcoholic beverages, including beer, wine, or spirits and/or mixed drinks made with spirits; |
| 12 | All types of savory bread, including French bread, baguette, Italian bread, homemade bread, Syrian or Arabic bread, sandwich loaf, hot dog or hamburger buns, dinner rolls, cheese bread, croissant, and packaged toast bread; with and without added spreads (butter, margarine, jam, etc.). The breads may be white or whole wheat. |
| 13 | All types of sweet breads, including, *broa^[1]^* , honey bread, sweet croissants, donuts, or churros. The breads might be homemade, bought from a bakery/cake shop or packaged with a brand; |
| 14 | Tapioca or cornmeal couscous or tapioca/beiju, with and without added spreads (butter, margarine, jam, etc.); |
| 15 | Crackers, with and without added spreads (butter, margarine, jam, etc.). The crackers may be white or whole wheat; |
| 16 | Biscuits, cookies or Graham crackers, with and without toppings or fillings; |
| 17 | Breakfast cereal, including ready-to-eat and cooked varieties, such as oats, granola, “Corn Flakes”, “All-Bran” or “Nesfit”, consumed with and without milk, sugar, honey, or artificial sweetener; |
| 18 | Porridge, yellow corn pudding (Brazilian ‘*curau”*), white corn pudding (Brazilian “*mungunzá”*); |
| 19 | Fresh fruits, including fruit salad, banana, orange or tangerine/mandarin, apple, papaya, mango, watermelon or cantaloupe, acai (either ready-to-eat or homemade from fruit or pulp), and any other fruit; |
| 20 | Dried fruits, peanuts, nuts or chestnuts, including salted, sugared or plain versions. |
| 21 | Cereal bar; |
| 22 | Soups, including homemade, ready-to-eat (frozen or canned) and instant (packaged) varieties. It includes chicken soup, vegetable soup (with and without noodles and meat), green broth, bean, lentil, or pea soup (with and without noodles or meat), ”*tacacá”^[2]^* , and any other type of soup; |
| 23 | Instant noodles; |
| 24 | Pasta, such as spaghetti, fettuccine, penne, or other types, including homemade or branded ready-to-eat/heat meals. Sauce options include simple tomato sauce, Bolognese sauce, bechamel or cheese sauce, and any other type of sauce. Sauces can be homemade or ready-to-eat or heat, such as tetra pack or sachet versions; |
| 25 | Lasagna, including homemade or branded ready-to-eat/heat meals, with a variety of sauces and fillings ( meat, ham, vegetable, or mixed); |
| 26 | Filled (or stuffed) pasta, such as ravioli, cannelloni, capeletti, and other types, including homemade or branded ready-to-eat/heat meals. Sauce varieties include classic tomato sauce, Bolognese sauce, Alfredo or cheese sauce, and any other type of sauce. Sauces can be homemade or ready-to-eat or heat, bag-in-box and pouch packed; |
| 27 | Savory pie recipes, including homemade or branded ready-to-eat/heat varieties. Filling options include chicken, meat, palm heart or other vegetables, and cheese; |
| 28 | Pizza, including homemade or branded ready-to-eat/heat varieties. Pizza toppings include mozzarella, margherita, Calabrese, pepperoni, Portuguese, ham, bacon, or any other flavor (such as pork loin, chicken, four-cheese, arugula, vegetarian, etc.); |
| 29 | Beans, lentils, or chickpeas, with and without meat, including mixed dishes such as feijoada and “*baiao de dois*”^[3]^; |
| 30 | Rice, including plain rice and rice-based preparations such as risotto. Rice may be white or whole grain; |
| 31 | French fries, which may have been homemade from scratch or prepared with frozen potatoes; |
| 32 | White potatoes or sweet potatoes, including mashed potatoes and other recipes; |
| 33 | Cassava/yucca, taro or yam, which may have been consumed boiled, fried, mashed or as part of other recipes; |
| 34 | Cassava flour, “pirão”^[4]^ , or farofa, which may have been prepared at home/restaurant or purchased ready-to-eat.; |
| 35 | Corn on the cob or canned corn; |
| 36 | Corn flour or “*farofa”*^[5]^, polenta, creamed corn, “*paulista couscous*”^[6]^ , Moroccan couscous, savory or sweet “*pamonha*”^[7]^ ; |
| 37 | Dark-green leafy vegetables, including lettuce, Swiss chard, watercress, arugula, kale, cabbage, spinach, endive, escarole, chicory, and others; |
| 38 | Vegetables, including tomato, cucumber, onion, pumpkin, zucchini, eggplant, carrot, beetroot, broccoli, cauliflower, green beans, chayote, bitter melon, okra, sweet pepper, radish, or other vegetables. These include all fresh, frozen, and dried options in cooked or raw forms. |
| 39 | Fresh or canned palm heart, fresh or canned mushrooms, and canned peas; |
| 40 | Soybeans or processed soy products and soy protein-based products, including tofu, steak and hamburger, meatballs; |
| 41 | Hamburger, chicken nuggets, and chicken tenders which may have been purchased in their branded or frozen forms or consumed at fast-food restaurants; |
| 42 | Dried beef, jerked beef, or salted meat, bacon, pork rinds, sausage and “*linguica*”^[8]^; |
| 43 | Fresh meats in general, including different cuts of beef, pork, poultry, and offal. Meats may have been raw/marinated, cooked, sautéed or stewed, roasted, grilled, barbecued, fried, breaded or prepared in forms; |
| 44 | Sushi, sashimi or temaki; |
| 45 | Fish in general, including fresh fish such as salmon, tuna, sardine, hake and tilapia, and fish that has been cured, or preserved, with salt, such as salted cod or canned fish. Fish may have been raw/marinated, cooked, sautéed or stewed, roasted, grilled, barbecued, fried, breaded or prepared in other forms; |
| 46 | Seafood in general, including fresh, dried and salted shrimp, crab, squid and octopus. Seafood may have been raw/marinated, cooked, sautéed or stewed, roasted, grilled, barbecued, fried, breaded or prepared in other forms; |
| 47 | Ham, mortadella, salami, turkey breast or other luncheon meats; |
| 48 | Chicken or other bird eggs, including fried, scrambled, boiled and eggs consumed as omelet or in other egg-based preparations; |
| 49 | All types of cheese, including mozzarella, fresh cheese, ricotta, cottage cheese, semi-cured cheese, cured cheese, rennet cheese, grated cheese, and other types of cheese; |
| 50 | Sweet or salty popcorn, which may be homemade popcorn (made from scratch using popcorn kernels) or industrialized popcorn (such as packaged or microwave popcorn); |
| 51 | Fried or baked savory snacks, including “*pastel*”^[9]^ “*coxinha*”^[10]^ , patty, esfiha, kibbeh, “*acarajé*” or “*abará*”^[11]^ and other savory snacks. Snacks may be homemade or purchased from full-service restaurant meals, ready-to-eat from grocery stores, or non-ready-to-eat from grocery stores; |
| 52 | Packaged chips or potato chips/straws, including “Cheetos”, “Doritos”, “Elma Chips”, “Ruffles” and other packaged chips; |
| 53 | Dressings in general, including ketchup or barbecue sauce, mayonnaise, mustard, soy sauce and salad dressings; |
| 54 | Cake with and without icing and/or filling, pies, panettone or “*cuca*”^[12]^ . These include homemade (from cake mix or made from scratch using flour, eggs and other culinary ingredients), or purchased ready-to-eat from grocery stores (such as Bauducco, Pullman, Ana Maria, and other brands) or non-ready-to-eat from grocery stores; |
| 55 | Ice cream, popsicle or ice pop; |
| 56 | Desserts in general, including rice pudding, coconut sweet, pudding, “*manjar*”^[13]^ or mousse, “*brigadeiro*”^[14]^ , gelatin, fudge, fruit preserves, guava paste, pumpkin or quince sweet, and other types of dessert. These include homemade desserts (from dessert mix or made from scratch), or purchased ready-to-eat from grocery stores (such as Bauducco, Pullman, Ana Maria, and other brands) or non-ready-to-eat from grocery stores; |
| 57 | Chocolate bars or candies, caramel or other types of candies, lollipop or chewing gum, peanut brittle, “*paçoca*”^[15]^ or peanut candy, cotton candy, and other sweets treats. |

**Supplementary Table S2. Comparison of dietary energy contribution of Nova food groups and subgroups estimated from the Nova24h and an interviewer-led 24 h recall (n=186)**

|  |  | % of total energy intake | | | | | | |  | Intraclass correlation coefficients | | |
| --- | --- | --- | --- | --- | --- | --- | --- | --- | --- | --- | --- | --- |
| Nova food groups and subgroups |  | Nova24h | | |  | Reference tool | | |  |  |  |  |
|  |  | Mean | 95% CI | |  | Mean | 95% CI | |  | ICC | 95% CI | |
| **Unprocessed or minimally processed foods** |  | **52.3** | **49.9** | **54.7** |  | **52.6** | **50.0** | **55.3** |  | **0.78** | **0.71** | **0.84** |
| Red meat |  | 8.4 | 6.8 | 10.0 |  | 7.0 | 5.5 | 8.5 |  | 0.83 | 0.77 | 0.87 |
| Milk and plain yogurt^1^ |  | 4.6 | 3.7 | 5.4 |  | 6.5 | 4.9 | 8.1 |  | 0.50 | 0.33 | 0.62 |
| Fruit |  | 7.8 | 6.7 | 8.8 |  | 5.7 | 4.8 | 6.6 |  | 0.78 | 0.67 | 0.85 |
| Grains |  | 5.5 | 4.8 | 6.3 |  | 5.4 | 4.4 | 6.4 |  | 0.67 | 0.56 | 0.75 |
| Legumes |  | 4.2 | 3.5 | 4.9 |  | 4.6 | 3.8 | 5.5 |  | 0.74 | 0.65 | 0.81 |
| Poultry |  | 4.0 | 3.1 | 4.9 |  | 3.9 | 3.0 | 4.8 |  | 0.76 | 0.68 | 0.82 |
| Eggs |  | 3.0 | 2.4 | 3.6 |  | 3.0 | 2.2 | 3.7 |  | 0.88 | 0.84 | 0.91 |
| Vegetables |  | 1.7 | 1.5 | 1.9 |  | 2.7 | 2.2 | 3.1 |  | 0.54 | 0.36 | 0.66 |
| Flours^2^ |  | 2.2 | 1.6 | 2.7 |  | 2.4 | 1.6 | 3.2 |  | 0.31 | 0.08 | 0.49 |
| Roots and tubers |  | 1.9 | 1.4 | 2.3 |  | 2.4 | 1.6 | 3.1 |  | 0.62 | 0.49 | 0.71 |
| Freshly squeezed fruit juices^3^ |  | 2.4 | 1.8 | 3.0 |  | 2.3 | 1.7 | 3.0 |  | 0.66 | 0.54 | 0.74 |
| Pasta |  | 2.5 | 1.6 | 3.4 |  | 2.0 | 1.2 | 2.7 |  | 0.85 | 0.80 | 0.89 |
| Fish and seafood |  | 1.5 | 0.8 | 2.2 |  | 1.2 | 0.6 | 1.8 |  | 0.80 | 0.73 | 0.85 |
| Nuts and seeds without salt, sugar, or oil |  | 1.2 | 0.8 | 1.7 |  | 1.1 | 0.5 | 1.5 |  | 0.69 | 0.59 | 0.77 |
| Coffee and tea |  | 0.6 | 0.5 | 0.6 |  | 0.9 | 0.8 | 1.1 |  | 0.50 | 0.31 | 0.64 |
| Other unprocessed or minimally processed foods^4^ |  | 0.8 | 0.3 | 1.2 |  | 1.5 | 0.9 | 2.1 |  | 0.43 | 0.24 | 0.57 |
| **Processed culinary ingredients** |  | **11.6** | **10.4** | **12.8** |  | **11.9** | **10.6** | **13.2** |  | **0.54** | **0.38** | **0.65** |
| Plant oils |  | 3.9 | 3.5 | 4.3 |  | 6.0 | 5.2 | 6.7 |  | 0.31 | 0.08 | 0.48 |
| Sugar, honey, or molasses |  | 4.2 | 3.5 | 4.8 |  | 2.9 | 2.2 | 3.6 |  | 0.67 | 0.55 | 0.75 |
| Animal fats^5^ |  | 1.8 | 1.4 | 2.3 |  | 2.0 | 1.3 | 2.7 |  | 0.65 | 0.53 | 0.74 |
| Other processed culinary ingredients^6^ |  | 1.7 | 1.1 | 2.3 |  | 1.0 | 0.4 | 1.5 |  | 0.58 | 0.44 | 0.68 |
| **Processed foods** |  | **17.1** | **15.2** | **19.0** |  | **14.7** | **12.5** | **16.5** |  | **0.72** | **0.62** | **0.79** |
| Cheese |  | 6.6 | 5.3 | 7.8 |  | 4.7 | 3.6 | 5.6 |  | 0.67 | 0.55 | 0.76 |
| Processed bread |  | 3.9 | 3.0 | 4.8 |  | 4.1 | 3.1 | 5.0 |  | 0.78 | 0.70 | 0.83 |
| Wine and beer |  | 2.9 | 1.8 | 4.0 |  | 2.4 | 1.2 | 3.6 |  | 0.84 | 0.78 | 0.88 |
| Processed cakes and desserts^7^ |  | 1.7 | 1.0 | 2.4 |  | 1.1 | 0.5 | 1.5 |  | 0.55 | 0.40 | 0.66 |
| Ham and other salted, smoked, or canned meat or fish |  | 0.8 | 0.4 | 1.2 |  | 0.9 | 0.5 | 1.4 |  | 0.70 | 0.61 | 0.78 |
| Nuts and seeds with salt or sugar |  | 0.5 | 0.2 | 0.9 |  | 0.8 | 0.1 | 1.5 |  | 0.47 | 0.30 | 0.61 |
| Savory snacks, including croquettes, pastries, and mini pies^8^ |  | 0.5 | 0.0 | 1.0 |  | 0.3 | 0.0 | 0.6 |  | 0.68 | 0.57 | 0.76 |
| Other processed foods^9^ |  | 0.2 | 0.1 | 0.3 |  | 0.5 | 0.3 | 0.8 |  | 0.20 | -0.06 | 0.40 |
| **Ultra-processed foods** |  | **19.0** | **17.0** | **20.9** |  | **20.8** | **18.5** | **23.2** |  | **0.75** | **0.66** | **0.81** |
| Cakes, cookies, and pies |  | 1.6 | 1.0 | 2.1 |  | 2.9 | 1.8 | 3.9 |  | 0.63 | 0.51 | 0.73 |
| Breads^10^ |  | 2.5 | 1.8 | 3.2 |  | 2.8 | 2.0 | 3.7 |  | 0.70 | 0.59 | 0.77 |
| Desserts and sweets^11^ |  | 2.6 | 1.9 | 3.3 |  | 2.8 | 1.9 | 3.6 |  | 0.72 | 0.63 | 0.79 |
| Reconstituted meat or fish products |  | 2.6 | 1.8 | 3.3 |  | 2.2 | 1.5 | 2.8 |  | 0.52 | 0.36 | 0.64 |
| Ice cream and ice pops |  | 1.8 | 1.0 | 2.5 |  | 1.3 | 0.6 | 2.0 |  | 0.82 | 0.76 | 0.86 |
| Crackers |  | 0.8 | 0.6 | 1.1 |  | 1.2 | 0.6 | 1.8 |  | 0.54 | 0.39 | 0.66 |
| Soft drinks |  | 1.1 | 0.6 | 1.6 |  | 1.2 | 0.7 | 1.7 |  | 0.83 | 0.78 | 0.88 |
| Milk-based drinks and flavored yogurts |  | 0.8 | 0.5 | 1.0 |  | 1.0 | 0.7 | 1.4 |  | 0.55 | 0.40 | 0.66 |
| Sauces, dressings, and gravies |  | 0.7 | 0.4 | 0.9 |  | 0.8 | 0.4 | 1.2 |  | 0.60 | 0.47 | 0.70 |
| Breakfast cereals |  | 0.5 | 0.3 | 0.7 |  | 0.8 | 0.3 | 1.2 |  | 0.54 | 0.38 | 0.65 |
| Margarine |  | 0.3 | 0.1 | 0.5 |  | 0.6 | 0.3 | 0.9 |  | 0.54 | 0.38 | 0.65 |
| Salty snacks |  | 0.4 | 0.1 | 0.6 |  | 0.3 | 0.0 | 0.6 |  | 0.78 | 0.71 | 0.83 |
| Instant and canned soups |  | 0.2 | 0.0 | 0.4 |  | 0.1 | 0.0 | 0.3 |  | 0.79 | 0.72 | 0.84 |
| Other ultra-processed foods^12^ |  | 3.1 | 2.1 | 4.1 |  | 2.8 | 1.9 | 3.6 |  | 0.30 | 0.07 | 0.48 |
| **Total** |  | **100.0** |  |  |  | **100.0** |  |  |  |  |  |  |

ICC, Intraclass correlation coefficients

^1^Includes milk consumed with coffee, fruit, and powdered chocolate

^2^Includes wheat flour, cassava flour and corn flour

^3^Includes juice from fruit pulp

^4^Includes a few homemade pastries, sweet or savory

^5^Includes salted and unsalted butter

^6^Includes textured soy protein, baking powder; vinegar; cornstarch

^7^Includes processed cakes and desserts (store-bought, prepacked for direct sale such as from deli counters)

^8^Includes deep-fried croquettes or pastries with different fillings (cheese, chicken, beef, ham, and others), and mini pies with a filling

^9^Includes processed juices and fruits and veggies preserved in brine

^10^Includes industrial-processed breads and buns

^11^Includes chocolate bars, candies, cereal bars and gelatin desserts, puddings, and *brigadeiro* (dessert made of condensed milk and cocoa powder)

^12^Includes soy protein-based drinks, fruit juices and energy drinks; other sweetened drinks; distilled alcoholic drinks; sweeteners; spread cheese; microwavable popcorn; frozen meals and ready-to-eat/heat pizza, savory pies, and French fries

1. Traditional Brazilian biscuit made with a base of corn flour.
2. A typical Brazilian soup made with dried shrimps, tucupí (wild cassava byproduct), alfavaca (wild Amazonian basil), manioc starch, hot yellow peppers, and jambú – a leafy plant with anesthetic properties.
3. A typical Brazilian recipe made with a base of rice and beans.
4. A typical Brazilian recipe made with a base of cassava flour.
5. Buttery and toasty Brazilian side dish made with cassava flour.
6. A typical Brazilian recipe made with a base of corn and cassava flour.
7. A typical Brazilian recipe made with a base of corn flour.
8. Brazilian-style smoked sausage.
9. A Brazilian style of fried pastry with fillings.
10. A Brazilian style of breaded pastry with a chicken filling.
11. Acarajé and abará are typical Brazilian dishes made with black-eyed peas filled with shrimp. Acarajé is shaped into balls and deep-fried in boiling *azeite de dende* also known as Brazilian palm oil.
12. A typical Brazilian cake with fruits or other fillings.
13. A typical Brazilian pudding made with a base of coconut.
14. A typical Brazilian dessert made of condensed milk, cocoa powder, butter, and chocolate sprinkles covering the outside layer.
15. A typical Brazilian peanut based sweet.
