## Supplemental Tables for "A novel web-based 24-hour dietary recall tool in line with the Nova food processing classification: description and evaluation"

| **Supplementary Table S3. Dietary contribution (% of total energy intake) of Nova food groups using the Nova24h tool** | | | | | | | | | | | |
| --- | --- | --- | --- | --- | --- | --- | --- | --- | --- | --- | --- |
| **or the interviewer-led 24-hour dietary recall (reference method) according to sex (n=186)** | | | | | | | |  |  |  |  |
| **Men (n= 83)** |  |  |  |  |  |  |  |  |  |  |  |
|  | Nova24h | | |  | Reference method | | |  | Intraclass correlation coefficients | | |
| Nova food groups | Mean | 95% CI | |  | Mean | 95% CI | |  | ICC | 95% CI | |
| Unprocessed or minimally processed foods | 50.7 | 47.3 | 54.0 |  | 50.8 | 47.0 | 54.5 |  | 0.8 | 0.7 | 0.9 |
| Processed culinary ingredients | 12.1 | 10.3 | 13.8 |  | 12.3 | 10.4 | 14.3 |  | 0.4 | 0.1 | 0.6 |
| Processed foods | 17.4 | 15.1 | 19.8 |  | 16.5 | 13.3 | 19.7 |  | 0.7 | 0.6 | 0.8 |
| Ultra-processed foods | 19.9 | 16.7 | 23.0 |  | 20.4 | 16.8 | 24.0 |  | 0.8 | 0.6 | 0.8 |
| **Women (n= 103)** |  |  |  |  |  |  |  |  |  |  |  |
|  | Nova24h | | |  | Reference method | | |  | Intraclass correlation coefficients | | |
| Nova food groups | Mean | 95% CI | |  | Mean | 95% CI | |  | ICC | 95% CI | |
| Unprocessed or minimally processed foods | 53.6 | 50.3 | 57.0 |  | 54.0 | 50.3 | 57.7 |  | 0.8 | 0.7 | 0.9 |
| Processed culinary ingredients | 11.3 | 9.6 | 12.9 |  | 11.6 | 9.8 | 13.3 |  | 0.6 | 0.4 | 0.7 |
| Processed foods | 16.8 | 14.0 | 19.7 |  | 13.2 | 10.6 | 15.8 |  | 0.7 | 0.6 | 0.8 |
| Ultra-processed foods | 18.3 | 15.7 | 20.8 |  | 21.2 | 18.1 | 24.3 |  | 0.7 | 0.6 | 0.8 |

| **Supplementary Table S4. Dietary contribution (% of total energy intake) of Nova food groups using the Nova24h tool** | | | | | | | | | | | |
| --- | --- | --- | --- | --- | --- | --- | --- | --- | --- | --- | --- |
| **or the interviewer-led 24-hour dietary recall (reference method) according to age group (n=186)** | | | | | | | | |  |  |  |
| **<40years (n=92)** |  |  |  |  |  |  |  |  |  |  |  |
|  | Nova24h | | |  | Reference method | | |  | Intraclass correlation coefficients | | |
| Nova food groups | Mean |  | 95% CI |  | Mean | 95% CI | |  | ICC | 95% CI | |
| Unprocessed or minimally processed foods | 53.2 | 49.8 | 56.6 |  | 53.8 | 49.9 | 57.7 |  | 0.8 | 0.7 | 0.8 |
| Processed culinary ingredients | 12.3 | 10.6 | 14.1 |  | 11.9 | 9.9 | 13.9 |  | 0.6 | 0.4 | 0.7 |
| Processed foods | 16.3 | 13.6 | 19.1 |  | 13.9 | 11.0 | 16.8 |  | 0.7 | 0.5 | 0.8 |
| Ultra-processed foods | 18.1 | 15.4 | 20.9 |  | 20.4 | 17.1 | 23.8 |  | 0.7 | 0.5 | 0.8 |
| **>= 40years (n=94)** |  |  |  |  |  |  |  |  |  |  |  |
|  | Nova24h | | |  | Reference method | | |  | Intraclass correlation coefficients | | |
| Nova food groups | Mean |  | 95% CI |  | Mean | 95% CI | |  | ICC | 95% CI | |
| Unprocessed or minimally processed foods | 51.4 | 48.1 | 54.8 |  | 51.4 | 47.7 | 55.0 |  | 0.8 | 0.7 | 0.9 |
| Processed culinary ingredients | 10.9 | 9.3 | 12.5 |  | 11.9 | 10.2 | 13.6 |  | 0.5 | 0.2 | 0.6 |
| Processed foods | 17.9 | 15.2 | 20.5 |  | 15.5 | 12.6 | 18.3 |  | 0.8 | 0.6 | 0.8 |
| Ultra-processed foods | 19.8 | 17.0 | 22.7 |  | 21.2 | 17.9 | 24.5 |  | 0.8 | 0.7 | 0.9 |

| **Supplementary Table S5. Dietary contribution (% of total energy intake) of Nova food groups using the Nova24h tool** | | | | | | | | | | | |
| --- | --- | --- | --- | --- | --- | --- | --- | --- | --- | --- | --- |
| **or the interviewer-led 24-hour dietary recall (reference method) according to education (n=186)** | | | | | | | |  |  |  |  |
| **Less than college/university (n=95)** |  |  |  |  |  |  |  |  |  |  |  |
|  | Nova24h |  |  |  | Reference method | | |  | Intraclass correlation coefficients | | |
| Nova food groups | Mean | 95% CI | |  | Mean | 95% CI | |  | ICC | 95% CI | |
| Unprocessed or minimally processed foods | 53.2 | 50.1 | 56.4 |  | 55.0 | 51.2 | 58.8 |  | 0.8 | 0.6 | 0.8 |
| Processed culinary ingredients | 12.0 | 10.2 | 13.7 |  | 11.6 | 9.7 | 13.5 |  | 0.5 | 0.3 | 0.7 |
| Processed foods | 17.0 | 14.3 | 19.8 |  | 14.1 | 11.4 | 16.7 |  | 0.7 | 0.6 | 0.8 |
| Ultra-processed foods | 17.8 | 15.2 | 20.4 |  | 19.3 | 16.1 | 22.5 |  | 0.8 | 0.7 | 0.9 |
| **College/university (n=91)** |  |  |  |  |  |  |  |  |  |  |  |
|  | Nova24h |  |  |  | Reference method | | |  | Intraclass correlation coefficients | | |
| Nova food groups | Mean | 95% CI | |  | Mean | 95% CI | |  | ICC | 95% CI | |
| Unprocessed or minimally processed foods | 51.3 | 47.8 | 54.9 |  | 19.3 | 16.1 | 22.5 |  | 0.8 | 0.7 | 0.9 |
| Processed culinary ingredients | 11.3 | 9.6 | 12.9 |  | 12.2 | 10.4 | 14.0 |  | 0.5 | 0.3 | 0.7 |
| Processed foods | 17.2 | 14.6 | 19.8 |  | 15.3 | 12.2 | 18.4 |  | 0.7 | 0.5 | 0.8 |
| Ultra-processed foods | 20.2 | 17.3 | 23.2 |  | 22.4 | 19.0 | 25.9 |  | 0.7 | 0.5 | 0.8 |

| **Supplementary Table S6. Dietary contribution (% of total energy intake) of Nova food groups using the Nova24h tool or the interviewer-led 24-hour dietary recall (reference method) according to weight status (n=186)** | | | | | | | | | | |  |
| --- | --- | --- | --- | --- | --- | --- | --- | --- | --- | --- | --- |
| **Not overweight/obese (n= 85)** |  |  |  |  |  |  |  |  |  |  |  |
|  | Nova24h | | |  | Reference method | | |  | Intraclass correlation coefficients | | |
| Nova food groups | Mean | 95% CI | |  | Mean | 95% CI | |  | ICC | 95% CI | |
| Unprocessed or minimally processed foods | 54.6 | 50.9 | 58.3 |  | 54.5 | 50.1 | 59.0 |  | 0.8 | 0.7 | 0.9 |
| Processed culinary ingredients | 11.3 | 9.5 | 13.0 |  | 11.5 | 9.5 | 13.6 |  | 0.5 | 0.3 | 0.7 |
| Processed foods | 16.8 | 13.8 | 19.9 |  | 14.9 | 11.5 | 18.3 |  | 0.8 | 0.6 | 0.8 |
| Ultra-processed foods | 17.3 | 14.7 | 19.9 |  | 19.0 | 15.6 | 22.5 |  | 0.7 | 0.5 | 0.8 |
| **Overweight/obese (n=101)** |  |  |  |  |  |  |  |  |  |  |  |
|  | Nova24h | | |  | Reference method | | |  | Intraclass correlation coefficients | | |
| Nova food groups | Mean | 95% CI | |  | Mean | 95% CI | |  | ICC | 95% CI | |
| Unprocessed or minimally processed foods | 50.4 | 47.3 | 53.4 |  | 50.9 | 47.8 | 54.0 |  | 0.7 | 0.6 | 0.8 |
| Processed culinary ingredients | 11.9 | 10.3 | 13.6 |  | 11.5 | 9.5 | 13.6 |  | 0.5 | 0.3 | 0.7 |
| Processed foods | 17.4 | 15.0 | 19.7 |  | 14.9 | 11.5 | 18.3 |  | 0.7 | 0.5 | 0.8 |
| Ultra-processed foods | 20.4 | 17.5 | 23.3 |  | 19.0 | 15.6 | 22.5 |  | 0.8 | 0.7 | 0.9 |
